# Frailty and mortality in hospitalized older adults with COVID-19: retrospective observational study

**DOI:** 10.1101/2020.05.26.20113480

**Authors:** Robert De Smet, Bea Mellaerts, Hannelore Vandewinckele, Peter Lybeert, Eric Frans, Sara Ombelet, Wim Lemahieu, Rolf Symons, Erwin Ho, Johan Frans, Annick Smismans, Michaël R. Laurent

## Abstract

**Background:** Older adults with coronavirus disease 2019 (COVID-19) face an increased risk of adverse health outcomes including mortality. Ethical guidelines consider allocation of limited resources based on likelihood of survival, frailty, co-morbidities and age. However, the association of frailty with clinical outcomes in older COVID-19 patients remains unclear.

**Objectives:** To determine the association between frailty and short-term mortality in older adults hospitalized for COVID-19.

**Design:** Retrospective single-center observational study.

**Setting and participants:** N = 81 patients with COVID-19 confirmed by reverse-transcriptase polymerase chain reaction (RT-PCR), at the Geriatrics department of Imelda general hospital, Belgium.

**Measurements:** Frailty was graded according to the Rockwood Clinical Frailty Scale (CFS). Demographic, biochemical and radiological variables, co-morbidities, symptoms and treatment were extracted from electronic medical records.

**Results:** Participants (N = 48 women, 59%) had a median age of 85 years (range 65-97 years), median CFS score of 7 (range 2 - 9), and 42 (52%) were long-term care residents. Within six weeks, eighteen patients died. Mortality was significantly but weakly associated with age (Spearman r = 0.241, *P* = 0.03) and CFS score (r = 0.282, *P* = 0.011), baseline lactate dehydrogenase (LDH) (r = 0.301, *P* = 0.009), lymphocyte count (r = -0.262, *P* = 0.02) and RT-PCR Ct value (r = -0.285, *P* = 0.015). Mortality was not associated with long-term care residence, dementia, delirium or polypharmacy. In multivariable logistic regression analyses, CFS, LDH and RT-PCR Ct values (but not age) remained independently associated with mortality. Both age and frailty had poor specificity to predict survival. A multivariable model combining age, CFS, LDH and viral load significantly predicted survival.

**Conclusions and implications:** Although their prognosis is worse, even the oldest and most severely frail patients may benefit from hospitalization for COVID-19, if sufficient resources are available.

**BRIEF SUMMARY:** Outcomes of frail older adults hospitalized for COVID-19, particularly long-term care residents, remain unclear. In this retrospective cohort, frailty predicted mortality independently of age or established biomarkers.

## INTRODUCTION

Coronavirus disease 2019 (COVID-19) is a global pandemic caused by severe acute respiratory syndrome coronavirus 2 (SARS-CoV-2).^1^ Older adults are at increased risk of hospitalization and mortality due to COVID-19.^2-5^

Different ethical guidelines deal with triage in case a surge in hospital admissions due to COVID-19 overwhelms scarce hospital resources.^6-8^ Likelihood of benefit, age and frailty are among the most commonly used triage criteria.^9,10^ In the U.K. and in Belgium (among other countries), intensive care unit (ICU) admission is not recommended for frail older adults aged 65 years and older.^11,12^ These guidelines rely on frailty assessment according to the Rockwood Clinical Frailty Scale (CFS). Patients can be classified on the CFS as not frail (scores 1-4), mildly frail (score 5), moderately frail (score 6) or severely frail (score 7-9).^13^ ICU admission is discouraged for frail older adults *i.e*. those with a CFS score of 5 or higher in the U.K. and Belgium.^11,12^ Hospital admission is discouraged for nursing home residents with suspected or confirmed COVID-19 and a CFS score of 7 or higher.^12^

Previous studies have shown that frailty is associated with worse outcomes in hospitalized older adults.^13,14^ However, little is known about the outcomes in frail older adults or long-term care residents hospitalized for COVID-19. Therefore, the aim of this retrospective observational study was to describe outcomes in hospitalized geriatric COVID-19 patients according to their age, degree of frailty and place of residence.

## METHODS

### Study design

A retrospective, single-center observational study was performed among COVID-19 patients at the Geriatrics Department of Imelda General Hospital in Bonheiden, Belgium, admitted between March 12^th^ and April 30^th^, 2020. Demographic, clinical, laboratory and radiographic parameters were extracted from electronic health records. Laboratory values included C-reactive protein (CRP, reference values < 5 mg/L), ferritin, D-dimers, lactate dehydrogenase (LDH), 25-hydroxyvitamin D levels and white blood cell, platelet and lymphocyte counts. Polypharmacy was defined as the use of five or more medications.

### Ethics

The Ethical Committee approved the research protocol and waived the need for informed consent, since it did not constitute a clinical study according to national and European regulations.

### Clinical procedures

COVID-19 was confirmed by reverse transcriptase polymerase chain reaction (RT-PCR) testing on nasopharyngeal swabs, using protocols validated within our national SARS-CoV-2 reference network.^15^ All patients admitted through the emergency department were screened for COVID-19 by low-dose chest computed tomography (CT). Findings on COVID-19 likelihood and extent of pulmonary involvement (CT-score ranging 0 - 25) were reported using a standardized radiological protocol as described previously.^15^

All COVID-19 patients in our hospital were hospitalized on dedicated wards under the care of a staff pulmonologist, nephrologist, infectious disease specialist or geriatrician, depending on their usual care team *(e.g*. nephrology in dialysis patients). Additional local criteria to admit patients under geriatric care were age 85 years or older, long-term care residence or equivalent home care *(i.e*. complete dependency on assistance for activities of daily living), patients with dementia or delirium, or patients aged 75 years and older with multiple co-morbidities and polypharmacy.

On admission, an experienced geriatrician scored premorbid frailty according to the CFS based on information from patients, their families, caregivers, primary care referral letters or long-term care records.

### Statistics

Results for continuous and categorical variables are reported as median and interquartile range (IQR), or number (percentage), respectively. Differences between survivors and non-survivors were examined using Mann-Whitney U-test and Chi-square test for continuous and categorical variables, respectively. Association of age, frailty and other baseline characteristics with mortality were evaluated by Spearman r and multiple logistic regression. Survival according to frailty status was examined using odds ratios, survival analyses (log-rank Mantel-Cox test) and receiver-operator curve (ROC) analysis. Two-tailed *P* values < 0.05 were considered significant. All analyses were performed using GraphPad Prism v8.4.2.

## RESULTS

Baseline characteristics of our cohort are shown in **Table** 1. Median age was 85 years (minimum 65, maximum 97 years), and 48 were women (59%). Median CFS score was 7 (range 2-9). Dementia had been diagnosed in 36 patients (44%), and 42 (52%) were long-term care residents. Polypharmacy was present in 52 (64%) subjects.

**Table 1.** Characteristics of the population according to survival status.

|  | **All (N=81)** | **Survivors**<br>**(N=62)** | **Non-survivors**<br>**(N=19)** | **P** | **Spearman**<br>**r** |
| --- | --- | --- | --- | --- | --- |
| **Sex** |  |  |  | 0.89 |  |
| Men | 33 (41%) | 25 (40%) | 8 |  |  |
| Women | 48 (59%) | 37 (60%) | 11 |  |  |
| **Age** | 85 (81-90) | 84.5 (79-89) | 88 (84-90) | 0.03 | 0.241 |
| **LTC resident** |  |  |  | 0.27 |  |
| Yes | 42 (52%) | 30 (48%) | 12 |  |  |
| No | 39 (48%) | 32 (52%) | 7 |  |  |
| **CFS score** | 7 (5-7) | 6 (4-7) | 7 (6-8) | 0.011 | 0.282 |
| **Dementia** |  |  |  | 0.77 |  |
| Yes | 36 (44%) | 26 (43%) | 10 |  |  |
| No | 45 (56%) | 36 (57%) | 9 |  |  |
| **Polypharmacy** |  |  |  | 0.52 |  |
| Yes | 52 (64%) | 41 (65%) | 11 |  |  |
| No | 29 (36%) | 21 (35%) | 8 |  |  |
| **CT-score** | 9 (5-12) | 9 (5-12) | 9.5 (5-16.5) | 0.39 |  |
| **PCR Ct value** | 24.65 (20.25-28.78) | 25.95 (21.50-29.38) | 21.65 (19.28-22.95) | 0.015 | -0.285 |
| **CRP (mg/L)** | 58 (32-89) | 56 (32-88) | 75 (39-130) | 0.20 |  |
| **CRP<sub>max</sub> (mg/L)** | 110 (59-155) | 88 (47-140) | 190 (110-270) | P<0.0001 | 0.453 |
| **LDH (U/L)** | 305 (235-396) | 286 (229-367) | 390 (315-493) | 0.009 | 0.301 |
| **Lymphocytes**<br>**(10<sup>3</sup>/μL)** | 0.9 (0.6-1.2) | 0.9 (0.7-1.25) | 0.55 (0.4-1.05) | 0.020 | -0.262 |
| **Lymphocyte**<br>**nadir (10<sup>3</sup>/μL)** | 0.7 (0.5-0.95) | 0.8 (0.6-1.1) | 0.4 (0.275-0.65) | 0.0001 | -0.419 |
| **Delirium** |  |  |  | 0.88 |  |
| Yes | 34 (42%) | 26 (42%) | 8 |  |  |
| No | 47 (58%) | 36 (58%) | 11 |  |  |
| **Length of stay**<br>**(days)** | 13 (8-18.5) | 13 (8-21) | 7 (3.75-15) | 0.050 |  |

Sex, place of residence, dementia, polypharmacy, extent of affected lung tissue on CT or CRP values at baseline did not differ between survivors and non-survivors. However, compared to survivors of COVID-19, non-survivors were significantly older (88.5 vs. 85 years, median age) and frailer (median CFS 7 vs. 6). Their PCR Ct values were also significantly lower (indicating higher viral load). Baseline LDH was significantly higher and baseline lymphocyte count lower in non-survivors. Baseline CRP, ferritin, D-dimer, white blood cell, platelet or 25-hydroxyvitamin D levels were not different (latter data not shown). Lymphopenia was present on admission in 48 patients (60%) and occurred during admission in 60 of 80 patients (75%; one patient was excluded due to chronic lymphocytic leukemia). The peak CRP and lymphocyte nadir reached during admission was higher among non-survivors, and these differences were highly significant. Length of stay tended to be shorter in those who died *(P =* 0.05).

Among these variables, the CFS score was associated with dementia *(P <* 0.0001, r = 0.602), long-term care residence *(P <* 0.0001, r = 0.465), and weakly with sex (lower frailty in males, *P =* 0.007, r = -0.296) and incident delirium *(P* = 0.043, r = 0.230). There was no significant association between CFS and older age in our cohort.

One out of seventeen patients died in the non-frail group (CFS score **1-4)**, compared to eighteen deaths among **64** frail patients, however this difference did not reach significance *(P =* **0.054). Figure 1A** shows survivors and non-survivors according to their age and CFS. Most deaths occurred in older, frailer patients. However, this group overlapped considerably with many surviving frail older patients. Kaplan-Meier curves also showed only a trend towards higher mortality in frail vs. non-frail subjects (Mantel-Cox log-rank *P =* **0.06, Fig. 1B)**.

**Figure 1.**
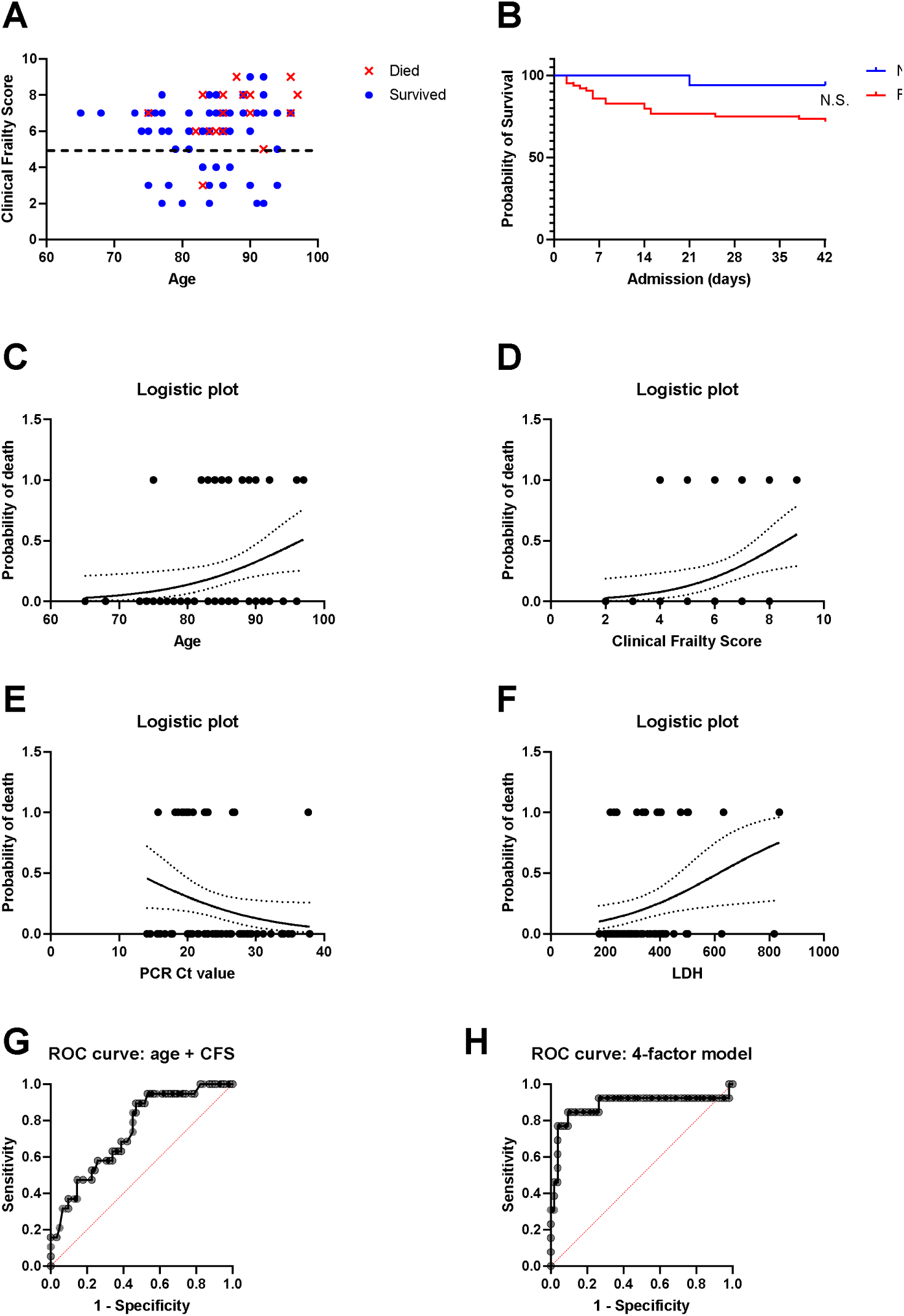
**(A)** Visual outline showing survivors and non-survivors according to age and CFS. **(B)** Kaplan-Meier curves for frail vs. non-frail individuals. **(C)** Probability of death from COVID-19 according to age, **(D)** CFS, **(E)** PCR Ct values, or **(F)** LDH in logistic regression models. **(G)** ROC curves for models with age + CFS, **(H)** age, CFS, PCR Ct values and LDH.

Next, we examined the clinical diagnostic utility of the individual variables that were significantly associated with mortality, in multiple logistic regression analyses. Again, age, CFS, PCR Ct values and LDH were significantly associated with higher odds of mortality **(Figure 1C-H** and **Table 2** below), whereas baseline lymphocyte count was no longer significant. In a bivariate model with age and CFS score combined, only the CFS remained significantly associated with mortality. The area under the ROC curve (AUROC) was 0.7443 (95% CI 0.6213-0.8673) for this model **(Figure 1G)**, with a positive and negative predictive value of 57% and 80%, respectively. When age and CFS were combined with PCR Ct values and LDH the latter three variables remained significantly associated with mortality. The AUROC for this model was 0.8824 (0.7384 - 1.000, *P <* 0.0001, **Figure 1H)**, with a negative predictive power of 89.5% and a positive predictive power of 78%, sensitivity of 54% and specificity of 78%.

**Table 2.** Odds ratios of four independent variables which predict mortality (outcome variable) in univariate and two multivariate models (logistic regression analyses).

|  | Single factor model | Model Age + CFS | Four-factor model |
| --- | --- | --- | --- |
| **Age** |  |  |  |
| OR | 1.117 (1.021-1.240)* | 1.096 (1.002-1.218) | 1.134 (1.002-1.333) |
| AUROC | 0.6638 (0.5311-0.7965)* |  |  |
| **CFS score** |  |  |  |
| OR | 1.705 (1.173-2.750)* | 1.588 (1.105-2.524)* | 1.752 (1.096-3.435)* |
| AUROC | 0.6876 (0.5547-0.8205)* |  |  |
| **PCR Ct value** |  |  |  |
| OR | 0.8969 (0.7980-0.9935)* |  | 0.8502 (0.7228-0.9718)* |
| AUROC | 0.6975 (0.5592-0.8359)* |  |  |
| **LDH (U/L)** |  |  |  |
| OR | 1.005 (1.001-1.010)* |  | 1.005 (1.000-1.011)* |
| AUROC | 0.7124 (0.5714-0.8534)* |  |  |

\* = $P < 0.05$

Seven patients were treated with hydroxychloroquine, 60 (74%) with antibiotics, 46 (57%) with i.v. fluid support and 25 with glucocorticoids (31%). Seven patients were admitted to ICU, five of whom died. The odds ratio for mortality was significantly higher in patients requiring ICU admission *(P =* 0.0017).

There were thirteen cases of presumably hospital-acquired COVID-19 (taking negative PCR on admission, incubation time and a local outbreak in one of our non-COVID-19 wards into account). Four of these patients died. There was no significantly higher or lower mortality between presumed hospital-acquired or community-acquired COVID-19 cases.

## DISCUSSION

The current COVID-19 pandemic particularly strikes frail older adults and/or long-term care residents, posing considerable medical and ethical challenges for overwhelmed healthcare systems. Different guidelines have been released to assist triage in this population.^9,11,16^ Belgian and U.K. guidelines recommend the CFS to inform decision making regarding hospital referral of nursing homes residents with suspected or confirmed COVID-19. However, empirical evidence supporting the use of frailty instruments to predict treatment outcomes and thus apply triage restrictions, has remained lacking.^17^

The short-term mortality (~23%) in this case series is similar to mortality rates reported for hospitalized older adults in Wuhan or California,^3,4^ but lower than reported by Sun *et al*.^18^ or than in the New York City area.^2^ This may be considered unexpected, given the greater frailty and older age of our patients compared to previous cohorts. Similar or higher mortality rates have been reported in long-term care residents^19^ or in younger ICU populations.^20^ These findings support the notion that it may be discriminatory and unethical to restrict hospital care based on age or frailty status alone.^10,21^ Still, mortality was higher in patients requiring ICU transfer in our cohort, suggesting that intensive care is of unclear clinical benefit in this population.^22^

Older age was significantly but weakly associated with increased risk of mortality, confirming recent studies.^1-4^ Anecdotally, nonagenarians or centenarians have survived COVID-19.^23^ Our main finding was that frailty was also significantly but weakly associated with higher risk of mortality in COVID-19 patients (multivariate odds ratio for mortality with each higher CFS point: 1.75.) Still, many severely frail patients survived (72%), and the CFS by itself had poor specificity and no useful cut-off for mortality prediction. A recent study from Italy showed that in N=105 COVID-19 patients, frailty as assessed by the fraity index was associated with in-hospital mortality or ICU admission, independent of age and sex.^24^

Apart from age and frailty, LDH was the only circulating biomarker significantly associated with mortality in our cohort. This confirms prior studies.^25,26,27^ However, only few patients met this criterion in our cohort, making it practically useless. Maximal CRP and nadir lymphocyte count during admission was significantly associated with mortality, but these parameters are not available at baseline. Interestingly, we observed a significant association between PCR Ct values and mortality. Viral load peaks longer in patients with more severe COVID-19 and in older adults, as shown by Zheng *et al*.^28^ We speculate that higher viral load may also be a marker for increased risk of mortality, although sampling bias needs to be excluded before we can support this conclusion. The four-factor model combining clinical, host and viral parameters showed the most promising characteristics, but still remained inadequate from a clinical perspective. Sun *et al*. reported a similar logistic regression model based on older age and lymphocyte count.^18^ Further work is needed to establish optimal clinical, viral and host immune system characteristics to predict mortality among COVID-19 patients.^26^

Our study provides the geriatric community with several novel insights into the outcomes of frail older COVID-19 patients. However, we recognize several limitations, mainly due to our retrospective study design. Since data were obtained retrospectively from electronic health records, missing data *(e.g*. for CT-scan or biochemical parameters) may have introduced bias, and follow-up was limited. However, selection bias is unlikely, since we included consecutive cases in a country with universal health coverage. Caution should be applied to extrapolate findings from this single-center study to other healthcare settings. The associations we observed may not be causally related. Despite our robust findings on the association between frailty and mortality, some analyses were likely underpowered due to our modest sample size. We chose not to include patients with so-called “radiographically confirmed” COVID-19 *i.e*. with typical clinical features and radiographic evidence on chest CT, but with repeatedly negative SARS-CoV-2 RT-PCR. However, only three such patients were excluded, which is unlikely to have influenced the results.

Many instruments to determine frailty are available.^29^ We applied the CFS, which has been adopted in several national COVID-19 triage policies, most notably by U.K. NICE guidelines.^11^ Previous research has shown that CFS scores can reliably be obtained in critically ill patients based on chart review, patient interview and/or family interview.^30^ However, we recommend further research to ascertain the reproducibility and reliability before widespread implementation of the CFS during COVID-19 outbreaks. Importantly, we were unable to include younger, non-frail patients, since frailty was not assessed in non-geriatric patients. The association between frailty and mortality would likely have been stronger if we included younger, less frail patients.

## CONCLUSIONS AND IMPLICATIONS

In summary, we showed that age and frailty were significantly but weakly associated with mortality among hospitalized older adults affected by COVID-19. However, both frailty and age alone have poor specificity to predict mortality, and many severely frail patients survived COVID-19. We recommend clinicians, ethicists and policy makers to consider these empirical findings.

## Data Availability

Underlying data will be made available to external researchers in good standing upon request.

## Author contributions

RDS and MRL designed the study, collected the data, analyzed the results and wrote the first draft. All authors contributed to the care of our COVID-19 patients, assisted in the data collection and analysis of the results, the writing of the manuscript and approved the final version.

